# Adverse Event Comparison between Glucagon-like Peptide-1 Receptor Agonists and Other Anti-Obesity Medications Following Bariatric Surgery

**DOI:** 10.1101/2024.04.21.24306138

**Authors:** Jason M Samuels, Kevin Niswender, Christianne L. Roumie, Wesley Self, Michael Holzman, Matthew Spann, C. Robb Flynn, Fei Ye, Rebecca Irlmeier, Luke M. Funk, Mayur Patel

## Abstract

**Introduction:** The safety of Glucagon-like Peptide 1 Receptor Agonists (GLP1RAs) following bariatric surgery remains largely unknown. This study aims to compare the incidence of adverse events (AEs) with GLP1RAs and other anti-obesity medications (AOMs) after bariatric surgery.

**Methods:** This single-center retrospective cohort included patients (ages 16-65) if they met the following criteria: underwent laparoscopic Roux-en-Y gastric bypass or sleeve gastrectomy (cohort entry date) and initiated AOMs. Subjects were categorized as users of ‘FDA-approved’ or ‘off label’ AOMs or GLP1RAs AOMs if documented as receiving the medication on the cohort entry date or after. Non-GLP1RA AOMs were Phentermine, Orlistat, Topiramate, Canagliflozin, Dapagliflozin, Empagliflozin, Naltrexone, Bupropion/Naltrexone, and Phentermine/Topiramate. GLP1RAs AOMs included: Semaglutide, Dulaglutide, Exenatide, Liraglutide. Primary outcome was AEs incidence. Logistic regression was used to determine the association of AOM exposure with AEs.

**Results:** We identified 599 patients meeting inclusion criteria (83% female, median 47.8 years old (IQR 40.9 – 55.4). Median surgery to AOM duration was 30 months (IQR 0 – 62). GLP1RAs were not associated with higher odds of AEs (adjusted Odds Ratio, (aOR) 1.1, [95% CI 0.5 – 2.6] and 1.1 [95% CI 0.6 – 2.3] for GLP1RA versus FDA-approved and ‘Off-Label’ AOMs, respectively. AOM initiation ≥12 months after surgery was associated with lower risk of AEs compared to <12 months (aOR 0.01. [95% CI 0.0 – 0.01], p<0.001).

**Conclusion:** GLP1RAs were not associated with an increased risk of AEs compared to non-GLP1RA AOMs in patients who previously underwent bariatric surgery. Prospective studies are needed to identify the optimal timeframe for initiation of GLP1RA.

## Introduction

Clinicians now have more options to treat obesity including the Glucagon-like Peptide 1 Receptor Agonists (GLP1RAs) than previously available. Medications such as Liraglutide, and more recently Semaglutide and the GLP1/GIP (glucose-dependent insulinotropic polypeptide receptor) receptor agonist Tirzepatide were initially developed for treatment of type 2 diabetes. The latest generation of GLP1RAs demonstrate significant weight loss, often achieving 15-20% weight loss.^1,2^ Older generations of anti-obesity medications (AOMs) such as Phentermine, Orlistat, and Topiramate remain the most commonly prescribed weight loss medications, but are rapidly being replaced.^3^

Bariatric surgery is the most effective treatment for severe obesity with over 50% of excess weight loss in most patients.^4^ Despite this, many patients do not achieve obesity resolution. More than 30% of patients regain weight after bariatric surgery,^5^ and 68% and 29% experience persistent severe obesity three years after sleeve gastrectomy and gastric bypass, respectively.^6^ Persistent severe obesity is especially common in patients with a preoperative Body Mass Index (BMI) >50 kg/m^2^, with more than half of patients failing to achieve obesity resolution.^7^ Clinicians require effective ‘adjuvant’ medical therapies for those who have previously undergone bariatric surgery.

The purpose of this study is to evaluate safety and weight loss of GLP1RA therapies compared to non-GLP1RA AOMs among patients who have previously undergone bariatric surgery. We further evaluated if there were differences in safety and weight loss among those who were prescribed medications before and after 12 months from the time of surgery.

## Methods

### Study Population

We assembled a retrospective cohort utilizing the Vanderbilt University Medical Center (VUMC) Synthetic Derivative, a de-identified database derived from the electronic health record for research purposes. Patients were included if they underwent laparoscopic Roux-en-Y gastric bypass (CPT code 43644, 43645) or laparoscopic sleeve gastrectomy (CPT 43775) prior to March 31, 2022. The surgical date was the cohort entry date. Further inclusion criteria were age 16-65 and BMI ≥35 kg/m^2^. All patients were included in the analytic cohort if they received a prescription for a GLP1RA AOM or non-GLP1RA AOM on or after the cohort entry date. Patients were excluded if they were prescribed GLP1RA or non-GLP1RA AOM prior to surgery. Patients were also excluded if they underwent revisional bariatric surgery. Follow-up of all patients was from the date of the initial prescription (index date) through either death, a discontinuation order in the electronic health record, or March 31, 2023 (study closure).

### Study Exposures: Non-GLP1 AOM or GLP1RA AOMs

Patients were categorized by the type of AOMs that documentation of use occurred on the index date (i.e. first documentation between time of surgery and end of data availability (March 31, 2023). FDA-Approved AOMs included: Phentermine, Orlistat, Bupropion/Naltrexone, and Phentermine/Topiramate. Off-label AOMS included: Topiramate, Canagliflozin, Dapagliflozin, Empagliflozin, Empagliflozin/Linagliptin, and Naltrexone. GLP1RA AOMs included: Semaglutide, Dulaglutide, Exenatide, and Liraglutide. If a patient received GLP1RA and non-GLP1RA exposures, then they were ineligible. Patient were allowed to cross over between FDA-Approved AOM and off-label AOMs, and if cross-over occurred, the patient was categorized based on the group in which the first prescription belonged. Similarly, patients were allowed to cross over between differing GLP1RAs. Metformin was not included.

### Outcomes

The co-primary outcomes were adverse events (AEs) and weight change. Serious adverse events (SAEs) were defined as any of the following: bowel obstruction, internal hernia, fistula of stomach, post-gastric surgery symptoms, complications of bariatric surgery, gastrojejunal ulcer, cholelithiasis/ cholecystitis, cholangitis/ choledocholithiasis, and GI hemorrhage. A complete list of AEs analyzed in this study are included in supplemental table 1 and included signs and symptoms commonly encountered after initiating a medication. Adverse events of interest were derived from previous publications of common adverse events following weight loss treatments.^8,9^ Analyses of weight changes utilized every measure of weight restricted to 12 months post-surgery through the end of follow-up and stratified by drug prescription.

### Covariates

Individual patient comorbidities at the time of cohort inclusion were collected from the VUMC Synthetic Derivative utilizing International Classification of Disease (ICD) 9/10 codes (supplementary table 1). Other covariates included surgery type (laparoscopic sleeve gastrectomy and laparoscopic Roux-en-Y gastric bypass), age (continuous), sex, and diabetes status.

### Statistical Analysis

Logistic regression models were used to determine the association of GLP1RA and FDA-approved AOM compared to off label AOM (reference) on SAEs and AEs accounting for comorbidities on the index date and the above covariates. To estimate the treatment effects over follow-up and to account for the time-dependent effect of weight on follow-up time, two interaction terms were included in the model: treatment by follow-up (months) and initial weight at the index date by follow-up time. Pairwise comparisons of 6-month and 12-month weights for patients prescribed non-GLP1RA AOMs and GLP1RAs AOMs were evaluated using means, and medians. A multivariable linear mixed-effects model was fit to determine the association of GLP1RA AOM compared to non-GLP1RA AOMs on weight loss. We evaluated the above outcomes stratified by whether the patient was prescribed the GLP1RA AOM or non-GLP1RA AOMs within 12 months of surgery or after 12 months. Statistical analyses were conducted using R version 4.3.2 (Vienna, Austria). This study was deemed exempt from institutional review by the VUMC Institutional Review Board (IRB #222066).This study follows the methodology and reporting recommendations of the Strengthening the Reporting of Observational Studies in Epidemiology (STROBE) guidelines for conducting observational cohort studies.^10^

## Results

### Cohort Demographics

The cohort identified 740 bariatric surgery patients who utilized a subsequent medication of interest. We excluded 141 patients (n=30 due to age, n=111 absence of baseline weight or BMI data, supplementary figure 1). Therefore, 599 patients were included in the analytic cohort. The exposure groups included off-label AOMs (n=266); GLP1RA AOM users (n=222) and FDA-approved AOMs (n=111). Baseline descriptive statistics can be found in Table 1. The cohort was 83% female, with a median age of 47.8 years (IQR 40.9 – 55.4), and 80% identified as white.

**Table 1:** Cohort Demographic Variables.

|  | Overall (%)<br>(N= 599) | GLP1RA<br>(N=222) | Legacy AOM<br>(N=111) | Off-Label<br>AOMs (N=266) |
| --- | --- | --- | --- | --- |
| **Age (years,**<br>**median (IQR))** | 47.8 (40.9 –<br>55.4) | 48.6 (42.6 -<br>56.1) | 47.4 (38.7 -<br>53.9) | 47.4 (40.8 -<br>55.6) |
| **Female** | 496 (83) | 173 (78) | 99 (89) | 224 (84) |
| **Race** |  |  |  |  |
| White | 478 (80) | 181 (82) | 88 (79) | 209 (79) |
| Black | 107 (18) | 32 (14) | 23 (21) | 52 (20) |
| Other | 10 (2) | 6 (3) | 0 (0) | 4 (2) |
| Missing | 4 (1) | 3 (1) | 0 (0) | 1 (0.4) |
| **Ethnicity** |  |  |  |  |
| Hispanic | 12 (2) | 5 (2) | 1 (1) | 6 (2) |
| Non-hispanic | 582 (97) | 216 (97) | 109 (98) | 257 (97) |
| Missing | 5 (1) | 1 (1) | 1 (1) | 3 (1) |
| **Surgery** |  |  |  |  |
| Gastric Bypass | 455 (76) | 152 (69) | 87 (78) | 216 (82) |
| Sleeve<br>Gastrectomy | 144 (24) | 70 (32) | 24 (22) | 50 (19) |
| **Comorbidities** |  |  |  |  |
| COPD | 137 (23) | 47 (21) | 20 (18) | 70 (26) |
| Asthma | 124 (21) | 42 (19) | 20 (18) | 62 (23) |
| Hypertension | 432 (72) | 176 (79) | 70 (63) | 186 (70) |
| Type 2 Diabetes | 282 (47) | 148 (67) | 28 (25) | 106 (40) |
| Osteoarthritis | 178 (30) | 64 (29) | 33 (30) | 81 (31) |
| Obstructive<br>Sleep Apnea | 373 (62) | 158 (71) | 66 (60) | 149 (56) |
| Stroke | 1 (0.2) | 1 (0.5) | 0 | 0 |
| Myocardial<br>Infarction | 6 (1) | 2 (1) | 1 (1) | 3 (1) |
| Hyperlipidemia | 298 (50) | 125 (56) | 40 (36) | 133 (50) |
| MASLD | 397 (66) | 169 (76) | 68 (61) | 160 (60) |
| GERD | 377 (63) | 137 (62) | 66 (60) | 174 (65) |
| Tobacco use | 15 (3) | 8 (4) | 3 (3) | 4 (2) |

The median time between surgery and drug initiation was 30 (IQR 0 – 62) months and baseline BMI (at drug initiation) was 39.2 kg/m^2^. Seventy-six percent underwent Roux-en-Y Gastric Bypass, with the remaining 24% having undergone sleeve gastrectomy. Comorbidities of the cohort were hypertension (72%), metabolic associated fatty liver disease (66%), gastroesophageal reflux disease (63%), and obstructive sleep apnea (62%). Follow-up time differed between the three exposure groups likely due to the later FDA approval of GLP1RAs compared to AOMs. Median follow-up was 48 months for the GLP1RA users versus 88 and 83 months for off-label and FDA-Approved AOM users, respectively (p<0.001).

### Serious and All Adverse Events

SAEs were rare (5.3%) across the entire cohort. On univariate analysis, the three groups did not differ in the incidence of all AE, or SAEs (p=0.9 for both, respectively). There was no difference in the risk of SAEs when comparing the off-label AOM exposure group to either the GLP1RA (p=0.74) or FDA-Approved AOM groups (p=0.88).

AEs including gastrointestinal complaints were common as 33% of patients experienced any adverse event with gastroesophageal reflux being the most common (25%). Abdominal pain was also frequent, reported in 18% of the full cohort (Table 2).

**Table 2:**
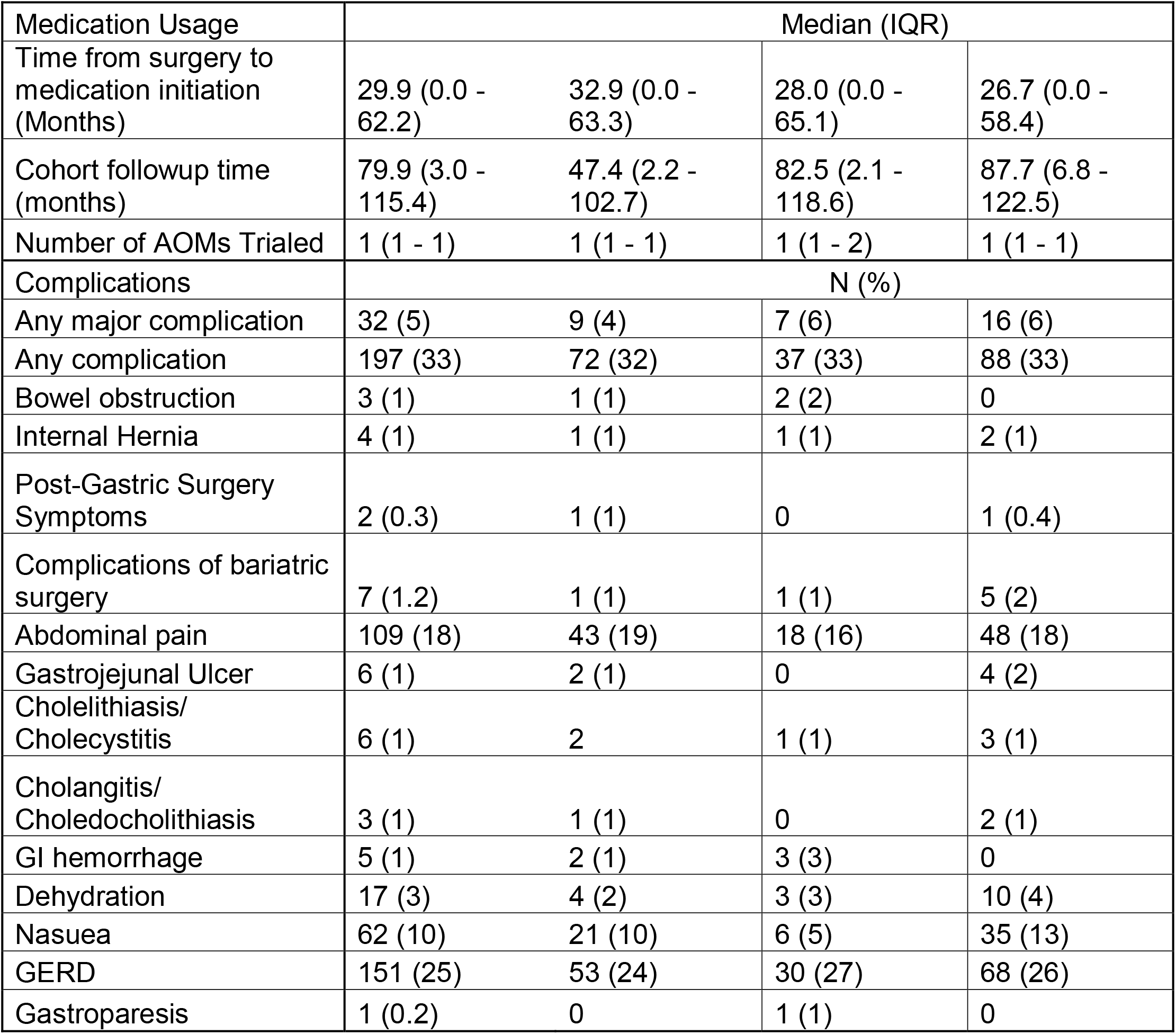
Incidence of Adverse Events with GLP1RA use after bariatric surgery.

Following adjustment for covariates, patients had a decreased odds of any AE if the AOM was initiated ≥12 months after surgery, compared to those who initiated any AOM <12 months after surgery (OR 0.01, 95% CI 0.0 – 0.01, p<0.001). Age, sex, type of surgery, or presence of diabetes was not associated with odds of AEs (Figure 1). There were insufficient episodes of AEs (n=8) to evaluate variables associated with these events in patients who initiated AOMs ≥ 12 months after surgery.

**Figure 1:**
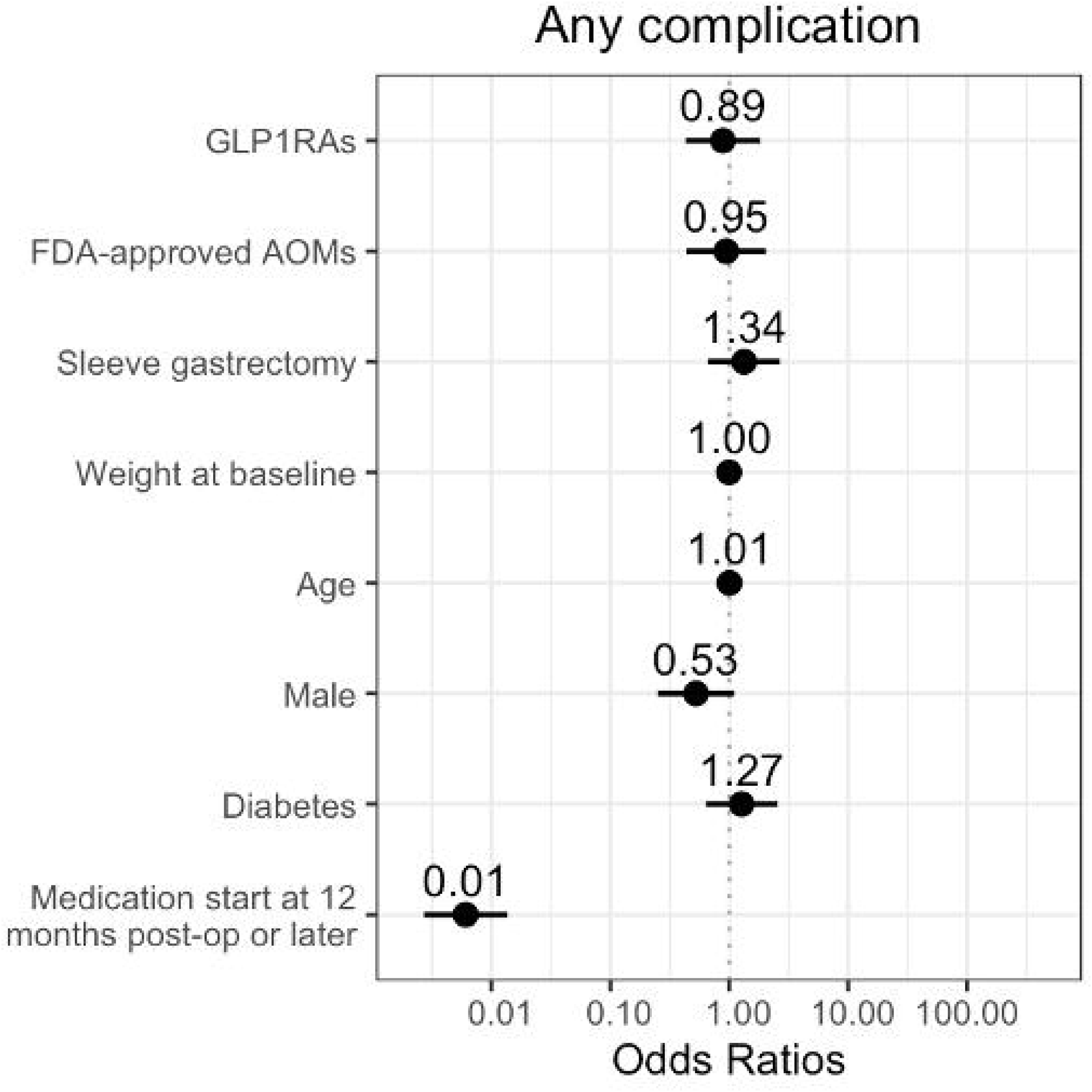
Forest Plot of variables associated with adverse events after bariatric surgery with off-label AOMs as the reference cohort.

### Changes in Weight

The cohort experienced total body weight (TBW) loss of 14% and 21% at six and twelve months after drug initiation, respectively. We separated the cohort into patients whose prescription medication was <12 months (n=246) and ≥ 12 months (n=353) after bariatric surgery. Among the subgroup who initiated AOMs ≥12 months after surgery, the GLP1RA AOM exposure was associated with the greatest TBW loss at 6 months compared to either FDA-approved or Off-label AOMs (−3.9% vs +0.9% and -1.9%, respectively p<0.001). However, at 12 months, this effect was diminished (−0.2% in GLP1RA AOM exposure group vs +4% and -0.9%, FDA-approved or Off-label AOMs respectively p<0.001).

Among the subgroups who initiated medications ≥12 months after surgery, we used a mixed-effects models with two interaction terms: medication exposure and duration (months), and baseline weight and time from drug start date to weight date. There was no difference in weight between off-label AOMs versus GLP1RA AOMs (p__interaction_=0.72) or with FDA-Approved AOMs (p__interaction_=0.91,). Male sex was associated with higher weight, with an expected 4.4 kg greater weight compared to females (95% CI 1.3 – 7.5, p=0.006). Age, sleeve gastrectomy, duration of AOM usage, the presence of diabetes, and medication duration were not significantly associated with weight loss (Figure 2). We further confirmed these findings by utilizing a linear regression for 12 months weights in the subgroup of those who initiated medications ≥12 months after surgery. In this analysis, we found that off-label AOM use was associated with a 4.6 kg lower weight compared to GLP1RA (95% CI 0.4 – 8.9 kg, p=0.03), and a 5.8 kg lower weight compared to FDA-approved AOMs (95% CI 0.5-11.1 kg, p=0.03) at 12 months. Age and sex remained associated with differences in weight as well, with a 2 kg decrease in weight for every 10-year increase in age (95% CI 0.3 – 4, p=0.03) and male sex associated with 6.8 kg greater weight (95% CI 0.5 – 13.2, p=0.03). Weight at medication prescription was associated with weight in the model, with a 0.9 kg greater weight for every 1 kg increase in baseline weight (95% CI 0.8 – 0.98, p<0.001).

**Figure 2:**
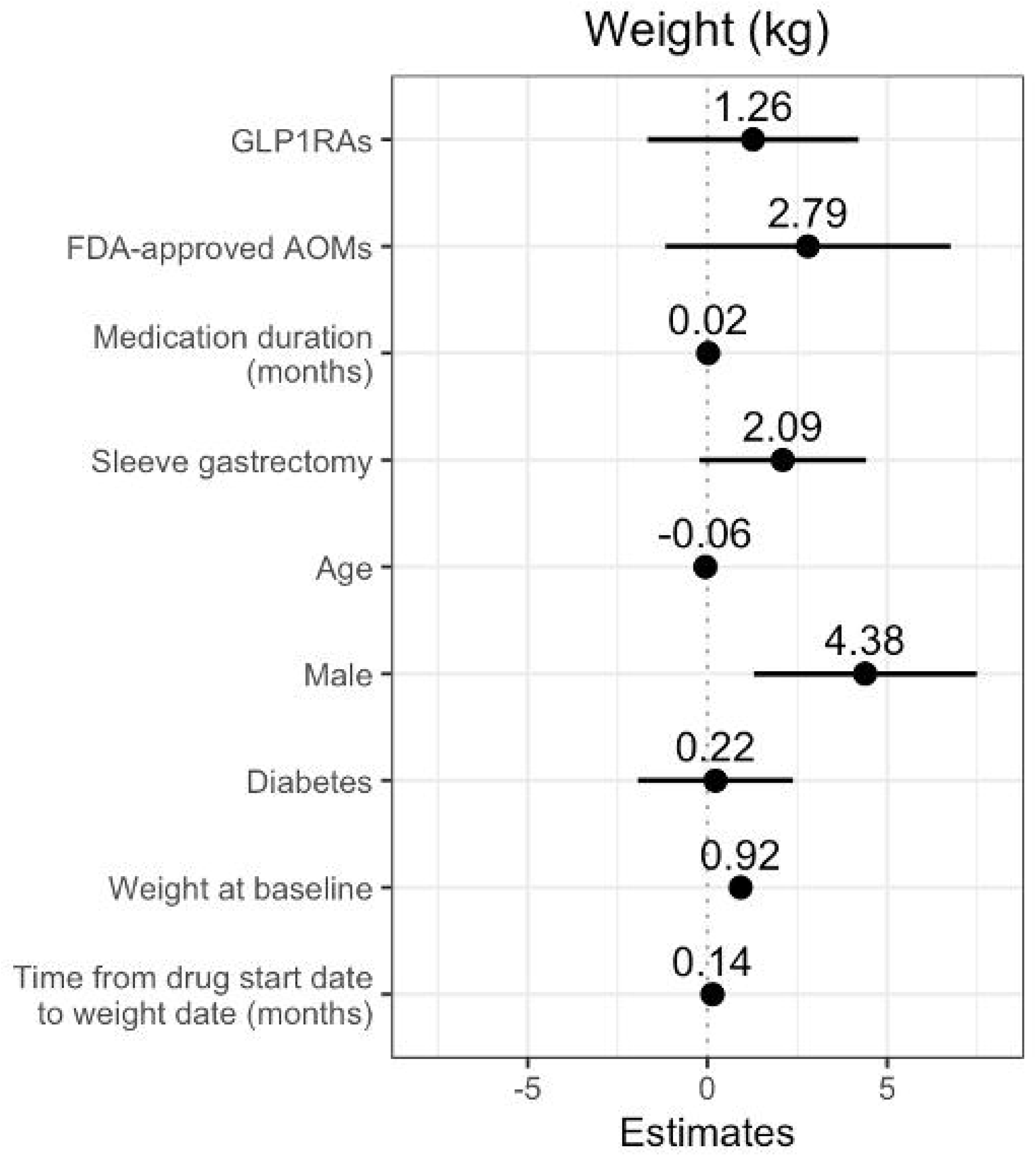
Forest Plot of effects of variables on weight for patients who initiated medications ≥ 12 months after bariatric surgery.

## Discussion

In this comparison of GLP1RA AOMs use with FDA-approved and off-label AOMs in postoperative bariatric surgery patients, we found that GLP1RA was associated with a similar risk profile of AEs. SAEs such as internal hernia, bowel obstructions, marginal ulceration, and gastrointestinal bleeding were rare in our cohort of patients, including those initiating GLP1RA within the first year of surgery. Overall, GLP1RAs appear safe in patients who have previously undergone bariatric surgery, but questions remain regarding the potential for enhancing weight loss with these medications.

A growing body of evidence supports the safety and feasibility of adjuvant medical therapy to enhance weight loss following bariatric surgery. Our study expanded upon prior findings by demonstrating GLP1RA therapies appear equivalently safe after bariatric surgery as non-GLP1RA AOMs, which have been widely used for weight regain or inadequate weight loss. Additionally, one-third of patients experienced an adverse event with GLP1RA therapy, which is in line with what has been described in the Semaglutide Treatment Effect in People With Obesity Trial (STEP) and the Study of Tirzepatide in Participants With Obesity or Overweight (SURMOUNT) trial in non-surgical patients.^11,12^ These findings align with prior studies which primarily reported mild to moderate GI side effects ranging in frequency from 30-50% with discontinuation rates of 5-9%.^13-16^ GLP1RA AOMs do not appear to lead to a heightened risk of adverse events in patients who have previously undergone bariatric surgery.

While our study identified an association between early (<12 month) initiation of any AOM after bariatric surgery and heightened risk of AEs, we cannot ascertain whether this association reflects the greater incidence of AEs of surgery itself during this period or a result of the addition of medications. Prior studies have identified a higher rate of AEs in the first 6 months after bariatric surgery, with few AEs occurring beyond 12 months.^8^ In our study, given that most complications occurred in the first 12 months (all but n=8 AEs occurred < 12 months after surgery), and that the three AOM exposure groups did not differ in the risk of AEs, we can theorize that this finding of greater AE risk reflects the heightened rate of AEs due to surgery rather than medication alone. This question ultimately requires prospective, interventional trials to ascertain the optimal timing for drug initiation and identify time at which the benefits and risk are optimally balanced.

While AOMs appear safe and feasible as a means to enhance weight loss after bariatric surgery, unanswered questions persist regarding how disease recurrence and suboptimal clinical response should be defined for patients who have undergone bariatric surgery.^17^ As a result, clinicians lack evidence-based practice guidelines for use of AOMs in patients who have previously undergone bariatric surgery. Few studies have investigated the impact of GLP1RA therapies on weight loss in patients who have undergone bariatric surgery, especially for newer GLP1RAs such as Semaglutide and Tirzepatide. To the authors’ knowledge, this is the largest study to date evaluating GLP1RA among patients who have previously undergone bariatric surgery.^18^ Two randomized trials investigating Liraglutide in patients with prior bariatric surgery demonstrated efficacy with 8% additional weight loss (compared to placebo), and liraglutide was well tolerated in these trials with no SAE.^16,19^ Two observational studies by Bonnett and Murvelashvili each reported that among patients with prior bariatric surgery, semaglutide can achieve 9 – 13% weight loss with a year of treatment.^20,21^ In order to realize the potential of adjuvant AOM, rigorous studies are needed to develop practice guidelines that direct the use of AOMs as adjuvant therapy. This includes clearly defining suboptimal response to bariatric surgery and obesity disease recurrence.

Our study has several limitations beyond the retrospective, observational nature of the study design. First, we cannot differentiate whether patients utilized these AOMs specifically for weight loss or for other clinical indications (i.e., topiramate for migraine prophylaxis or GLP1RAs for management of type 2 diabetes). Specifically, patients utilizing GLP1RA for weight loss require higher doses than that for diabetes control, and as we do not have dosing data, we cannot make this determination. We addressed this concern using two methods; first we excluded patients who utilized these agents prior to undergoing bariatric surgery, and second, we controlled for diabetes as a covariate. Given that 2/3^rd^ of our patients had a diagnosis of diabetes, this at least in part explains the lower weight loss achieved in our study compared to prior studies. Secondly, we cannot ascertain whether patients filled the prescriptions or took the medications, and there are likely instances in which patients were documented as utilizing these medications in the medical record, but due to shortages or prohibitive costs, were not actually receiving GLP1RA or other AOMs. Likewise, we are dependent upon accurate documentation in the medical record as to when these medications are initiated. Inaccurate date of prescribing or exposure misclassification may have contributed to the lower efficacy for weight loss. It is possible that patients previously initiated AOMs and had already reached a attained a weight loss nadir by the medication prescription date noted in the medical record. Finally, absent a comparator group who underwent surgery and were non users of AOMs, we cannot clearly attribute weight changes or AEs to the use of AOMs versus surgery alone. This salient limitation is most pertinent among the subgroup prescribed medications ≤1 year after surgery. However, the incidence of adverse events in our analysis coincides with those reported previously, with up to half of patients experiencing an adverse event in the years after bariatric surgery.^8^

## Conclusion

We report that use of GLP1RA as adjunctive therapy post-bariatric was not associated with increased risk of AEs compared to FDA approved and off label AOMs. This study also found minimal association with GLP1RA agents and weight change but is limited in determining a true impact on weight with GLP1RA in patients with a history of bariatric surgery absent specific dosing data. This study further highlights the promising role of GLP1RAs in augmenting weight loss after bariatric surgery. To fully realize this potential, clinical care requires evidence-based guidelines that define suboptimal response to bariatric surgery and obesity recurrence and optimal use including timing, indications for initiation, and long-term effects in this specific clinical context.

## Supporting information

STROBE Diagram

## Data Availability

All data produced in the present study are available upon reasonable request to the authors

