## Supplementary material for "Adverse Event Comparison between Glucagon-like Peptide-1 Receptor Agonists and Other Anti-Obesity Medications Following Bariatric Surgery": STROBE Diagram

### Slide 1
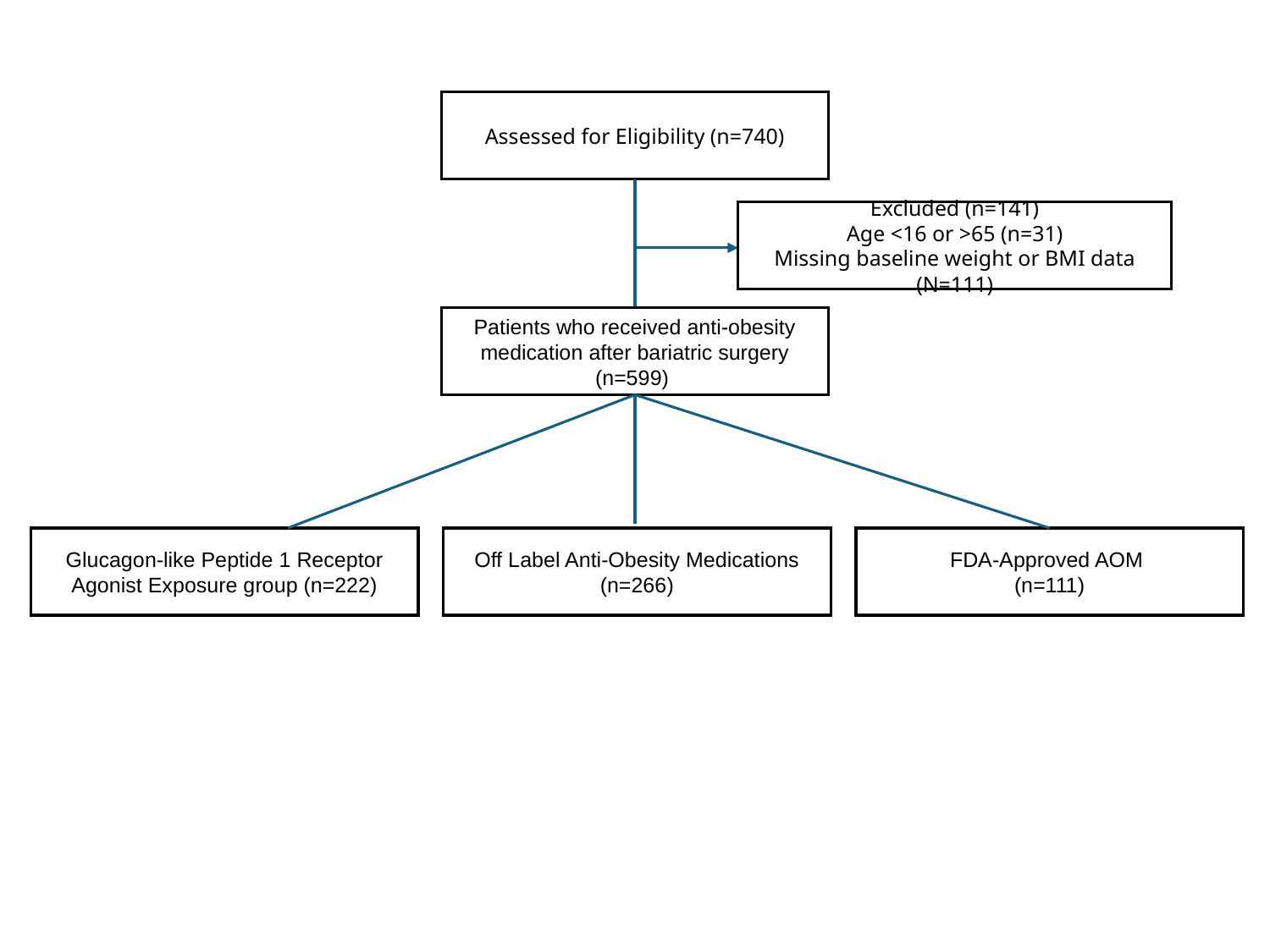

Assessed for Eligibility (n=740)
Excluded (n=141)
Age <16 or >65 (n=31)
Missing baseline weight or BMI data (N=111)
Patients who received anti-obesity medication after bariatric surgery (n=599)
Glucagon-like Peptide 1 Receptor Agonist Exposure group (n=222)
Off Label Anti-Obesity Medications (n=266)
FDA-Approved AOM
(n=111)
